# PD GENEration: An International Parkinson’s Disease Genetic Research Study

**DOI:** 10.64898/2026.05.20.26353696

**Authors:** Kamalini Ghosh Galvelis, Allison A. Dilliott, Megan Dini, Rebeca De León, Max Thom, Ignacio Azcarate, Nicola Bothwick, Lark Caboy, Anny Coral-Zambrano, Kirby Doshier, Megan Finke, Melissa Nicewaner, Sarah Osborne, Joshua Ruffner, Addison Yake, Adolfo Diaz, Tatiana Foroud, Anne Hall, Laura Heathers, Sarah Woody Lawrence, Karen Marder, Ignacio Mata, Niccolò E. Mencacci, Anna Naito, Martha Nance, John Poma, Ruth B. Schneider, Michael A. Schwarzschild, Tanya Simuni, Jennifer Verbrugge, Anne-Marie Wills, Yun Lu, Harry Gao, Ben Casavant, Cornelis Blauwendraat, Andrew B. Singleton, James C. Beck, Roy N. Alcalay, The Parkinson’s Foundation PD GENEration Study

## Abstract

**Background:** PD GENEration (NCT04057794, NCT04994015), sponsored by the Parkinson’s Foundation in partnership with Aligning Science Across Parkinson’s (ASAP) through the Global Parkinson’s Genetics Program (GP2), is an international, observational, clinical research study that offers genetic testing and counseling to people living with Parkinson’s disease (PwP) at no financial cost. PD GENEration has aimed to empower PwP and their clinicians with knowledge of their genetic status, to accelerate recruitment into precision medicine trials, and to advance research through data sharing. Since its launch in 2019, the study has expanded to enroll over 32,000 PwP (as of March 31, 2026), from 10 countries across North, Central, and South America, the Caribbean, and Israel.

**Methods:** Over the course of 6 years, PD GENEration has evolved to accommodate the growing scientific and research needs of the Parkinson’s community while also increasing the ability to return genetic test results to PwP at a greater scale. Participants with a diagnosis of Parkinson’s disease (PD) may enroll in-person or virtually where informed consent and blood sample collection can occur. Samples are analyzed at a College of American Pathologists/Clinical Laboratory Improvement Amendments (CAP/CLIA)-certified laboratory using whole genome sequencing, with variants curated for a primary panel of seven PD-associated genes. Results are disclosed during a genetic counseling visit, where further testing is offered for two optional additional gene panels. Those who consent undergo analysis of additional genes, and results are returned during a genetic counseling visit for those that test positive for a variant. In addition to returning genetic results to PwP, a central pillar of the study design has been the open sharing of genomic data to advance discovery in PD research in partnership with ASAP and GP2.

**Discussion:** PD GENEration applies a flexible framework, allowing for country specific considerations and the integration of multiple site models, evolving based on participant needs and the prioritization of equity and accessibility. We summarize PD GENEration’s implementation and scaling, highlight key accomplishments and lessons learned, and provide guidance for those interested in implementing large-scale clinical genetic testing studies across other diseases and therapeutic domains.

## Background

Although 10-15% of people diagnosed with Parkinson’s disease (PD) have been found to carry likely pathogenic or pathogenic variants associated with their disease [1], clinical genetic testing is not yet applied routinely in clinical care. Clinicians have historically been hesitant to implement these practices due to their limited impact on disease management and concerns regarding lack of insurance coverage for testing [2]; however, these views are beginning to change [3]. Enrollment into clinical trials has also become increasingly reliant on people living with PD (PwP) knowing whether their disease can be linked to genetic variants, with ongoing precision medicine clinical trials currently seeking to recruit individuals with genetic variants in *GBA1*, *LRRK2*, and *PRKN* [4–10]. PwP who have undergone clinical genetic testing have responded positively to their experience, regardless of their genetic status, with the majority reporting satisfaction with having information regarding potential disease risk that they could share with family members [11]. Yet, even with the recent general agreement that there are benefits to genetic testing in the PD landscape, costs and insufficient access to genetic counselors are still prohibitive to the testing process as part of routine clinical care.

To address this gap in testing, the Parkinson’s Foundation launched PD GENEration (NCT04057794, NCT04994015) an international, observational, clinical research study that offers genetic testing and counseling to PwP at no financial cost [1] in partnership with Aligning Science Across Parkinson’s (ASAP) through the Global Parkinson’s Genetics Program (GP2). Since it began in 2019, the study has expanded to enroll over 32,000 PwP (as of March 31, 2026), from 10 countries across North, Central, and South America, the Caribbean, and Israel.

The goals of PD GENEration are threefold: to empower PwP and their clinicians with knowledge of their genetic status, to accelerate recruitment into precision medicine trials, and to advance research through data sharing. This knowledge supports a better understanding of disease etiology, facilitates thoughtful discussions with family members and clinical care teams, and ultimately enables better informed decisions about clinical trial participation.

Here, we describe the implementation and evolution of PD GENEration at scale, including infrastructure changes over time, key accomplishments, challenges encountered, and lessons learned. We also highlight practical considerations that may inform development of similar large-scale clinical genetic testing studies across other diseases entities and therapeutic domains.

## Methods

### Study Design and Timeline

PD GENEration is a multi-center, cross-sectional study that has evolved as it has grown in reach. Here, details are provided on the different phases of the study, the current operational workflow as of March 2026, participant recruitment, engagement and retention, and budgetary considerations.

#### Pilot Phase: September 2019-December 2020

In September 2019, PD GENEration was launched by the Parkinson’s Foundation as a pilot study, offering – for the first time – College of American Pathologists/Clinical Laboratory Improvement Amendments (CAP/CLIA) certified genetic testing and genetic counseling to PwP, at no financial cost. The pilot aimed to assess feasibility of genetic testing and, through a randomized design, the experience of participants in relation to return of results either from their enrolling clinician or through a centralized genetic counseling resource, as previously described [12].

All participants were enrolled at one of six clinic-based sites of the Parkinson Study Group (PSG), a consortium of academic movement disorders specialists, and completed a set of questionnaires and scales to gather information regarding demographics, medication status, baseline knowledge regarding PD genetics, comorbid conditions, and family history of PD (**Table 1**). Clinical assessments included the Movement Disorder Society–Unified Parkinson’s Disease Rating Scale (MDS-UPDRS) [13] and the Montreal Cognitive Assessment (MoCA) [14]. Genetic analysis was performed using a clinical exome sequencing backbone covering approximately 4,700 genes [15]. However, variant curation was limited to seven PD-associated genes (*GBA1, LRRK2, PRKN, SNCA, PINK1, PARK7, VPS35*) (**Table 2**), defined based on a formal review of the landscape of genetic testing in PD at the time of study design [16, 17], and participants only received clinical genetic testing results for pathogenic or likely pathogenic variants. With a target enrollment of 600 participants, the Pilot Phase ultimately recruited 620 PwP, of which 525 completed genetic counseling and 327 completed outcome surveys [12]. Enrollment was temporarily paused in March 2020 due to the COVID-19 pandemic. During this period, the study protocol was adapted to support remote data collection and telemedicine-based assessments, allowing enrollment to resume by June 2020.

**Table 1.** Overview of enrollment goals, genetic testing methods, and data collection based on the PD GENEration study phase.

| PD GENERation Phase | Enrollment Goal (Cumulative) | CAP/CLIA Genetic Testing Method | Primary PD Gene Panel | Optional Secondary Panels | DNA Biobanking | Post-Test Genetic Counseling | Family History | Demographics | PD History | MDS-UPDRS | MoCA | Patient and Provider Surveys |
| --- | --- | --- | --- | --- | --- | --- | --- | --- | --- | --- | --- | --- |
| **Pilot** | 600 | CES | ✓ | - | ✓ | ✓ | ✓ | ✓ | ✓ | ✓ | ✓ | ✓ |
| **Clinical Phase** | 2000 | CES | ✓ | - | ✓ | ✓ | ✓ | ✓ | ✓ | ✓ | ✓ | ✓ |
| **Registry Phase: Clinical Exome** | 15,000 | CES | ✓ | - | ✓ | ✓ | ✓ | ✓ | ✓ | - | - | - |
| **Registry Phase: Whole Genome** | 30,000 | WGS | ✓ | ✓ | ✓ | ✓ | ✓ | ✓ | ✓ | - | - | - |

Abbreviations: CAP/CLIA, College of American Pathologists/Clinical Laboratory Improvement Amendments; CES, clinical exome sequencing; MDS-UPDRS, Movement Disorder Society-Sponsored Revision of the Unified Parkinson's Disease Rating Scale; MoCA, Montreal Cognitive Assessment; PD, Parkinson's disease; WGS, whole genome sequencing.

**Table 2.**
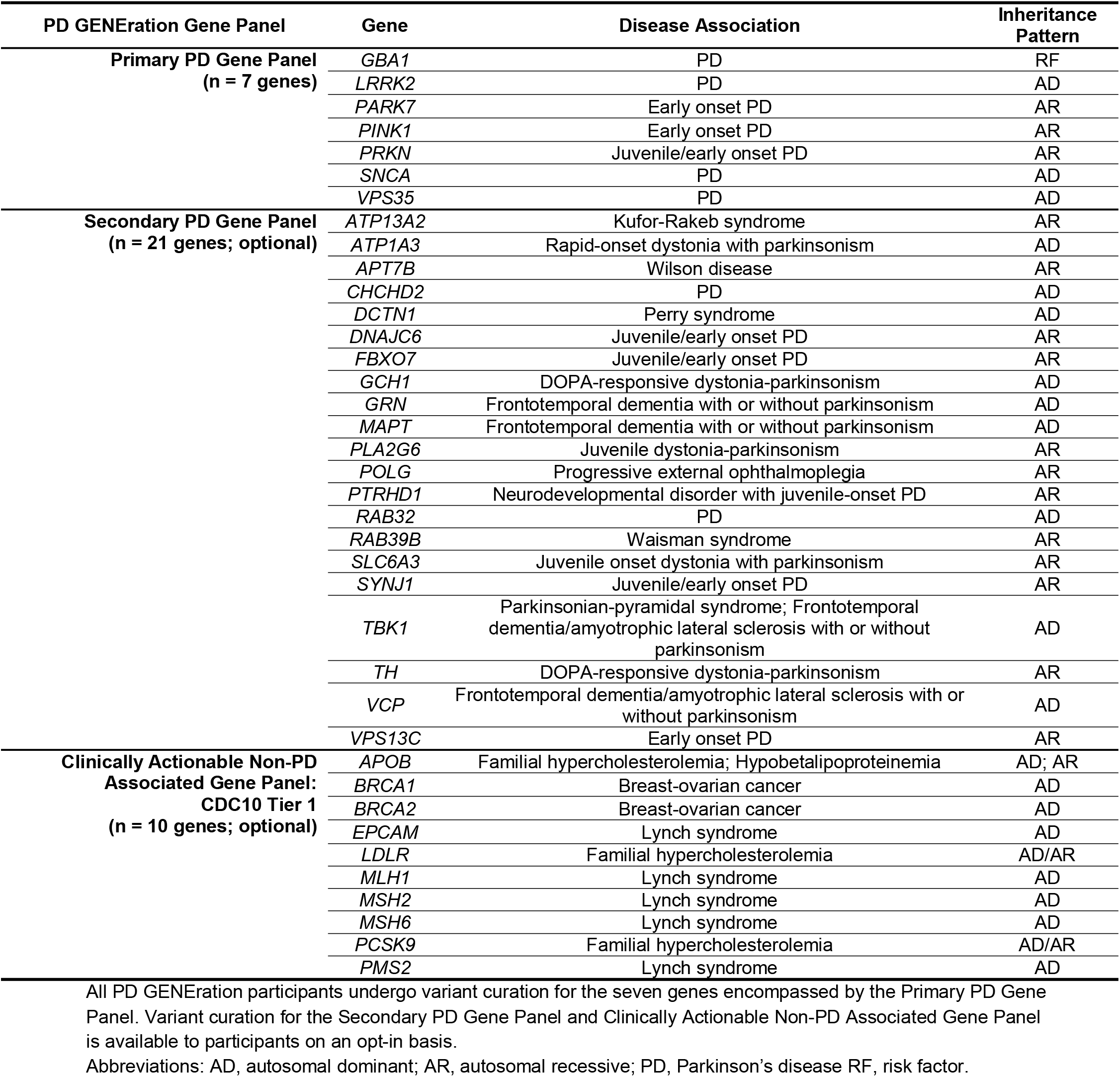
Genes included on the PD GENEration primary and secondary gene panels offered to participants through the study’s Registry Phase: Whole Genome Sequencing.

| PD GENERation Gene Panel | Gene | Disease Association | Inheritance Pattern |
| --- | --- | --- | --- |
| **Primary PD Gene Panel**<br>**(n = 7 genes)** | *GBA1* | PD | RF |
|  | *LRRK2* | PD | AD |
|  | *PARK7* | Early onset PD | AR |
|  | *PINK1* | Early onset PD | AR |
|  | *PRKN* | Juvenile/early onset PD | AR |
|  | *SNCA* | PD | AD |
|  | *VPS35* | PD | AD |
| **Secondary PD Gene Panel**<br>**(n = 21 genes; optional)** | *ATP13A2* | Kufor-Rakeb syndrome | AR |
|  | *ATP1A3* | Rapid-onset dystonia with parkinsonism | AD |
|  | *APT7B* | Wilson disease | AR |
|  | *CHCHD2* | PD | AD |
|  | *DCTN1* | Perry syndrome | AD |
|  | *DNAJC6* | Juvenile/early onset PD | AR |
|  | *FBXO7* | Juvenile/early onset PD | AR |
|  | *GCH1* | DOPA-responsive dystonia-parkinsonism | AD |
|  | *GRN* | Frontotemporal dementia with or without parkinsonism | AD |
|  | *MAPT* | Frontotemporal dementia with or without parkinsonism | AD |
|  | *PLA2G6* | Juvenile dystonia-parkinsonism | AR |
|  | *POLG* | Progressive external ophthalmoplegia | AR |
|  | *PTRHD1* | Neurodevelopmental disorder with juvenile-onset PD | AR |
|  | *RAB32* | PD | AD |
|  | *RAB39B* | Waisman syndrome | AR |
|  | *SLC6A3* | Juvenile onset dystonia with parkinsonism | AR |
|  | *SYNJ1* | Juvenile/early onset PD | AR |
|  | *TBK1* | Parkinsonian-pyramidal syndrome; Frontotemporal dementia/amyotrophic lateral sclerosis with or without parkinsonism | AD |
|  | *TH* | DOPA-responsive dystonia-parkinsonism | AR |
|  | *VCP* | Frontotemporal dementia/amyotrophic lateral sclerosis with or without parkinsonism | AD |
|  | *VPS13C* | Early onset PD | AR |
| **Clinically Actionable Non-PD**<br>**Associated Gene Panel:**<br>**CDC10 Tier 1**<br>**(n = 10 genes; optional)** | *APOB* | Familial hypercholesterolemia; Hypobetalipoproteinemia | AD; AR |
|  | *BRCA1* | Breast-ovarian cancer | AD |
|  | *BRCA2* | Breast-ovarian cancer | AD |
|  | *EPCAM* | Lynch syndrome | AD |
|  | *LDLR* | Familial hypercholesterolemia | AD/AR |
|  | *MLH1* | Lynch syndrome | AD |
|  | *MSH2* | Lynch syndrome | AD |
|  | *MSH6* | Lynch syndrome | AD |
|  | *PCSK9* | Familial hypercholesterolemia | AD/AR |
|  | *PMS2* | Lynch syndrome | AD |

All PD GENERation participants undergo variant curation for the seven genes encompassed by the Primary PD Gene Panel. Variant curation for the Secondary PD Gene Panel and Clinically Actionable Non-PD Associated Gene Panel is available to participants on an opt-in basis.
Abbreviations: AD, autosomal dominant; AR, autosomal recessive; PD, Parkinson's disease RF, risk factor.

Following Pilot Phase completion, participant satisfaction was assessed and >80% of those queried found utility in their results for many reasons, including satisfaction of participating in clinical research, better understanding of family risk, and fulfilling personal curiosity [11]. Interviews were also completed with the neurologists and genetic counselors that performed the return of results, which confirmed that PD GENEration was filling a gap in PD clinical care [18]. Based on the success of the Pilot, the study transitioned to its Clinical Phase, which focused on increased enrollment and continued data collection.

#### Clinical Phase (August 2020 - December 2021)

In the Clinical Phase, PD GENEration transitioned from a randomized pilot design to an expanded hybrid implementation model with 12 PSG movement disorder sites that launched enrollment in August 2020. Clinical data collection continued, with a greater reliance on telemedicine-based assessments to support broader participation (**Table 1**). In total, 1,966 participants were enrolled across the Pilot and Clinical Phases in the US, with 1,957 completing genetic testing and counseling.

The Pilot and Clinical Phases of PD GENEration demonstrated that: 1) PwP were interested in genetic testing and counseling and were satisfied with the process of results disclosure, whether it be from a PSG site clinician or centralized certified genetic counselor (CGC); and 2) the broad PD community, including PwP, clinicians, and researchers, recognized the value of accessible, large-scale genetic testing and counseling, highlighting the feasibility and acceptability of scalable genetic testing and counseling models in PD.

#### Clinical Exome Registry Phase (February 2021 - March 2025)

Although PD GENEration met its initial objectives of feasibility and participant satisfaction, the Parkinson’s Foundation had set a goal of enrolling 15,000 participants by December 2025 to ensure sufficient numbers of PwP were screened in the event of eligibility for potential precision medicine clinical trials. Feedback from participants and sites indicated that the collection of clinical data was a significant draw on time and created a backlog of PwP who wanted to participate. Moreover, the deep phenotyping conducted as part of PD GENEration would need to be repeated for future precision medicine trial inclusion and the consideration of the costs of infrastructure support for deep phenotypic data collection applied in the Pilot and Clinical Phases [1] ultimately led to the adoption of a simplified protocol (**Table 1**), with enrollment beginning in February 2021.

The objective of the Registry Phase was to significantly expand study accessibility through three recruitment models: local recruitment sites, nationally recruiting supersites, and referral sites, as further described in the Methods section. These models allowed for the expansion of the study to the entire US through more than 50 local PSG sites, inclusive of PSG sites; four national supersites; and more than 20 referral sites, as well as a pilot expansion to local recruitment sites in Canada and the Dominican Republic.

By the time the first Registry Phase concluded, a total of 16,003 PwP were enrolled in the study, with 15,169 having completed genetic testing and counseling. Of these, 189 PwP were enrolled in Canada and 314 were enrolled in the Dominican Republic. The Registry Phase of PD GENEration demonstrated strong engagement from the PD community, highlighting a willingness to participate in PD genetics research when accessible testing models are available, and delineated a clear mechanism by which the scalability of a large-scale genetic testing initiative could be achieved.

#### Whole Genome Sequencing Registry Phase (March 2024 - Present)

As the study evolved and with additional grant support, PD GENEration was able to expand in two distinct ways. First, PD GENEration transitioned to whole genome sequencing (WGS) to expand the scope of genomic data collected and shared with the research community. Second, the study made a concerted effort to expand internationally. In January 2024, PD GENEration formalized its ongoing partnership with GP2, a resource program of ASAP [19]. The purpose of this partnership was for the PD GENEration study to more directly contribute to GP2’s own effort to understand the complex genetic architecture underlying PD.

With the support of ASAP and GP2, the study revised its protocol and transitioned to WGS beginning in March 2024. Not only did this permit an order of magnitude increase in data that was shared with the research community but also created a compelling reason to share more genetic information back to participants beyond the initial 7-gene PD panel. These additional genetic tests include a panel of 21 secondary PD and parkinsonism-associated genes and a panel of ten clinically actionable genes associated with non-PD conditions (CDC10 Tier 1) (**Table 2**). Each of the additional genetic testing panels is optional and accompanied by additional genetic counseling sessions for participants that test positive for genetic variation.

Another goal of the partnership with GP2 was to broaden the contribution by those not typically represented in current PD genetic databases, which are largely composed of those of European ancestry. Here, PD GENEration not only increased outreach to underrepresented communities in the US but also built upon the success of the Dominican Republic and established a partnership with the Latin American Research consortium on the GEnetics of Parkinson’s Disease (LARGE-PD) study. These efforts further increased accessibility and PD GENEration expanded enrollment into additional countries across North, Central, and South America, the Caribbean, and Israel. The methodology applied in this current Registry Phase is described in detail below.

## Study oversight

PD GENEration is strategized and operationalized by the Parkinson’s Foundation’s clinical research team (PD GENEration team), which includes sub-teams focused on: US, Canada, and Israel operations, Latin America (LATAM) operations, regulatory affairs, genomics, data science and management, and community/patient engagement. Additionally, the study is supported by external vendors and partners including: 1) Navitas Clinical Research (Gaithersburg, MD), a full service contract research organization (CRO) for operations and data management; 2) Fulgent Genetics (El Monte, CA), a CAP/CLIA certified clinical genetic testing laboratory; 3) Indiana University School of Medicine (Indianapolis, IN), the centralized genetic counseling core; and 4) Tasso, Inc. (Seattle, WA), an at-home peripheral blood collection device company. The study is scientifically supported by GP2, a resource program of the ASAP initiative. The study meets regulatory approval through a central institutional review board (IRB), Advarra, Inc., and local site-based IRBs. Oversight is provided by a steering committee consisting of movement disorder specialists, PD genetic scientists, and PwP. There is also significant involvement from the Parkinson’s community, specifically from the Parkinson’s Foundation People with Parkinson’s Advisory Council (PPAC). PPAC consists of PwP and care partners who ensure the perspectives of PwP are integrated into Parkinson’s Foundation programs. For PD GENEration, PPAC has played an important part in each of the study’s phase transitions. PPAC members provide direct feedback on operational changes and are often among the first participants to engage in new study initiatives and sub-studies, offering practical insight into usability, participant experience, and feasibility.

## Study Workflow

All participants follow the same study workflow regardless of the recruitment model and location (**Figure 1**). Starting with a baseline visit, participants complete informed consent following site specific processes to: 1) undergo WGS, 2) receive genetic testing and counseling on the primary panel of seven PD-associated genes, and 3) be re-contacted regarding new results or additional research opportunities in the future. Participants are also given the option to provide preliminary consent to additional genetic testing, including a panel of 21 PD and parkinsonism-associated genes and/or the CDC10 Tier 1 panel.

**Figure 1.**
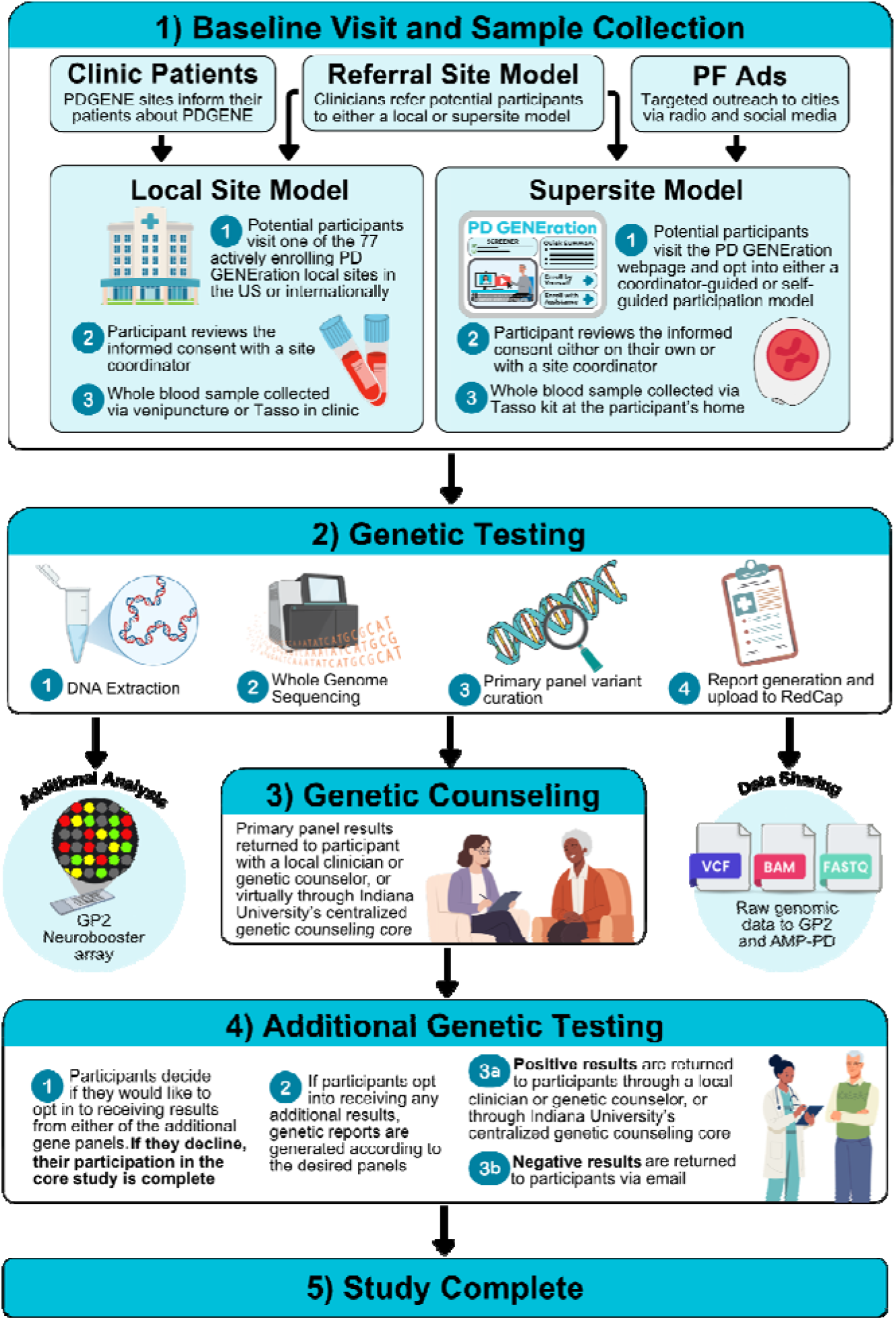
PD GENEration Registry Phase: Whole Genome Sequencing core study workflow. Participants are informed of the PD GENEration study through clinic visits, referral sites, Parkinson’s Foundation events, or advertisements and may enroll in-person or virtually through local sites or nationally enrolling supersites, where informed consent and blood sample collection occur. Samples are analyzed at a College of American Pathologists/Clinical Laboratory Improvement Amendments (CAP/CLIA)-certified laboratory using whole genome sequencing, with variants curated for the Primary Parkinson’s disease (PD) Gene Panel (n = 7 genes). Results are disclosed during a genetic counseling visit, where optional testing is offered for two additional gene panels. Those who opt in undergo analysis and positive genetic findings are disclosed through a second genetic counseling session. The workflow described is based on the current protocol as of March 31, 2026.

Following consent, participants watch a pre-test genetic counseling video and complete a survey within the electronic data capture (EDC) system to gather contact information, PD history, and family history of PD. A blood sample is collected either via venipuncture or utilizing a Tasso+ device—an FDA approved blood collection kit that enables at-home or site-assisted capillary blood collection from the upper arm without the need of a phlebotomist. The PD GENEration team has created study-specific instructional videos and materials for participants and care partners who may be using Tasso+ at home.

Once a blood sample is collected, it is shipped directly to the genetic testing lab for genomic analyses. Upon arrival, DNA is extracted from the blood sample and WGS is performed using the clinical genetic testing laboratory’s established methodologies and quality control processes.

Variants identified from WGS data within the seven PD-associated genes included in the primary PD GENEration panel are curated by clinical testing laboratory personnel using the American College of Medical Genetics and Genomics/Association for Molecular Pathology (ACMG/AMP) guidelines. At the request of PD GENEration, the laboratory also classified two variants within *GBA1* as “risk factor” variants, namely c.1093G>A p.Glu365Lys and c.1223C>T p.Thr408Met, as although they are not associated with the gene’s other associated phenotype, Gaucher disease, they are relevant as inclusion criteria for *GBA1*-targeted precision medicine trials for PD. Details regarding the sequencing, variant calling, and variant curation methodology, including quality control, were previously described [1]. If a participant is found to carry one or more variants classified as pathogenic, likely pathogenic, or a risk factor in any of the seven genes, a positive genetic report is issued describing all variant details, including the gene, genomic change, zygosity, and supporting evidence for pathogenicity. If no pathogenic, likely pathogenic, or risk factor variants are identified in the seven genes, a negative genetic report is issued. The report is securely returned to the study’s EDC system through an application programming interface (API) and is made available to site clinicians or the study’s centralized genetic counseling core. Typically, the clinical genetic testing laboratory returns the primary PD panel report within 4-6 weeks of sample arrival.

Genetic counseling sessions are conducted either in-person or virtually and include result disclosure and discussion of potential clinical and familial implications by site clinicians, site genetic counselors, or centralized CGCs. Centralized genetic counseling occurs via CGCs through a partnership with Indiana University’s genetic counseling core. Site clinicians have the option to either return results themselves or have results returned via the CGCs. Site clinicians who want to disclose results must first complete training consisting of videos and documentation that cover topics such as consent, pre-test discussion, genetic testing and reports, genetic counseling resources, results disclosure and post-test discussion, an example genetic counseling session for negative and positive results, and additional information on secondary health findings, created by the CGCs from Indiana University. Participants are also provided with gene-specific and family sharing fact sheets and receive a follow-up letter summarizing their results. Participants are encouraged to share their results with their clinicians if counseling is not performed by their primary PD medical provider. The duration of a genetic counseling session can vary person to person and is dependent on the results being returned (either negative or positive), family implications, clinical trial eligibility, and the number of questions the participant has [11]. Typically, based on review during the Pilot Phase of the study, a negative genetic counseling session takes between 10 minutes to 30 minutes and a positive session between 30 minutes to 2 hours. Once results are returned, participants confirm their previous consent decision and willingness to proceed with testing using the optional additional gene panels.

If the participant consents to one or both additional gene panels, the clinical genetic testing laboratory performs variant curation for the panels of interest in the same manner as for the primary panel. If found to be likely pathogenic or pathogenic variant positive, participants are scheduled for an additional genetic counseling session to review the results via a site clinician, site CGC, or centralized CGC.

Although the return of results concludes the participants’ initial engagement with the study, if new or updated genetic results are reported by the clinical testing laboratory based on variant re-curations, the participant is notified and another genetic counseling session is conducted. In parallel, all raw WGS data are returned to PD GENEration from the clinical genetic testing laboratory for downstream analysis.

## Participant Eligibility and Recruitment

PD GENEration considers any person with a confirmed diagnosis of PD who is 18 years or older eligible for enrollment. Full eligibility criteria are available in **Supplemental Table 1**. All participants provide informed consent and may choose to opt out of the study at any time.

Since its onset, PD GENEration has been a bilingual study, in English and Spanish. With all documents translated into Spanish, the study was able to offer testing and counseling capacities to Spanish-speaking populations in the United States and internationally, starting with expansion into Puerto Rico and the Dominican Republic in 2021, and throughout LATAM in 2024. Additionally, expansion into Israel in April 2024 prompted the addition of Hebrew as a third language and all participant-facing documents were translated and IRB approved. As of March 2026, the study is available to participants in the US and Puerto Rico, 7 countries throughout LATAM (Argentina, Chile, Colombia, El Salvador, Dominican Republic, Mexico, and Peru), Canada, and Israel.

PD GENEration recruitment relies on local recruitment sites, nationally recruiting supersites, and referral sites (**Figure 1**). Local sites may enroll their clinic-based population and PwP living in surrounding areas either in-clinic or virtually via a telemedicine visit. Nationally recruiting supersites allow eligible participants to enroll virtually via the Parkinson’s Foundation website or through telemedicine, regardless of where they receive their PD care. The supersite model, which is currently only active in the US, allows participants who may not have access to a local site the opportunity to participate in PD GENEration by enrolling online. The criteria to become a supersite are limited and entail the ability to enroll participants from anywhere in the US, based on IRB regulations, and have increased staffing capacity to support higher levels of enrollment and follow-up. As of March 2026, there are six active supersites staffed with clinicians, coordinators, and in most cases, CGC, who can facilitate participation of the entire study virtually. Of the six supersites, four have the capacity to enroll in both English or Spanish and two enroll in English only. Lastly, the referral site model allows for a more autonomous approach to supporting recruitment. Referral sites include medical and non-medical organizations that work to connect eligible participants to the PD GENEration study but do not conduct research, collect data, or enroll people with PD themselves. Rather, they share information about the study and help connect interested eligible participants to local sites or supersites, thereby allowing clinics that may not have staffing capacity or expertise to conduct the study to extend the opportunity for genetic testing and counseling to their patients.

### Community Outreach

Over the past six years, the PD GENEration team has utilized the robust existing footprint of the Parkinson’s Foundation to expand clinical research outreach efforts, increasing study awareness and accessibility. PD GENEration information is included in all Foundation presentation decks, literature, and event signage and scripts. The PD GENEration team has trained Foundation staff and volunteers in key talking points about the study, enabling meaningful conversations about how to enroll, why genetics matter to Parkinson’s, and how research advances treatments. Several internal teams at the Parkinson’s Foundation have aided the PD GENEration team and contributed to the success of PD GENEration’s expansion, including through operational and recruitment support.

PD GENEration also leverages the Parkinson’s Foundation’s strong community presence by informing and educating PwP about the study through its Global Care Network and at events, support groups, presentations, and other outreach activities. In 2021, the PD GENEration team began to partner with the Community Engagement and Events staff to host events in priority markets, promoting the availability of the study onsite, registering interest, following up with PwP, and finally, testing eligible participants onsite.

In addition to efforts in the United States, PD GENEration has conducted educational and recruitment-focused outreach events across LATAM in close collaboration with local sites and LARGE-PD. These initiatives have delivered culturally tailored education, expanded access to genetic testing resources, and strengthened community engagement, further supporting equitable participation in PD research.

## International expansion

Having all participant-facing materials and resources available in Spanish paved the way for expansion from the US into the Dominican Republic in 2021. This pilot served as an opportunity to learn about the unique needs of the country’s participants, assess infrastructure, and review the regulatory requirements. Recognizing that genetic return of results research was not previously available in the Dominican Republic, a more robust protocol was developed with additional clinical measures to generate a more comprehensive dataset, mirroring the Clinical Phase of PD GENEration. The community was highly receptive, with strong engagement from local and national regulatory bodies, clinicians, and participants. However, as seen with the clinical phase in the US, the logistical demands of the protocol created a level of burden that was not sustainable long-term, and the pilot concluded in 2023. Nevertheless, the Dominican Republic has since been reactivated with the streamlined Registry protocol that is employed as part of a broader expansion across LATAM.

With the lessons learned from the pilot in the Dominican Republic, a formal partnership was established with LARGE-PD to further expand the study into LATAM [20]. Starting with 5 sites, the partnership provided regional operational support and fostered shared trust with local communities, enabling a successful expansion. Each country presents distinct regulatory and logistical challenges, requiring protocol adaptations to support PD GENEration execution in LATAM. This included addressing challenges related to sample shipment and customs to ensure consistent delivery to the clinical testing laboratory in the US. Furthermore, CGCs are generally not available in LATAM. To coordinate return of results to participants, the PD GENEration team worked closely with CGCs at Indiana University School of Medicine to develop a comprehensive training program for clinicians who would be returning results in LATAM. The additional training and support have enabled LATAM clinicians to effectively perform all necessary aspects of the PD GENEration workflow independently. Following the success of the initial LATAM pilot, the study has expanded to a total of 12 sites in seven countries (**Supplemental Table 2**). The return of results training program has trained and certified 33 clinicians across 13 LATAM countries as of March 2026. These efforts enable PD GENEration to reach patients in rural areas as clinicians engage them during their existing community rounds.

In addition to expansion across LATAM, PD GENEration has also expanded to two sites in Canada and three sites in Israel. The expansions reflect the study’s broader international growth and continued efforts to increase global accessibility.

## Participant Retention and Engagement

To ensure that participants fully benefit from the study, PD GENEration strives to minimize the lost to follow-up (LTFU) rate. Common points of LTFU events have been observed at sample collection and scheduling of genetic counseling, likely reflecting challenges inherent to virtual models of enrollment. Similar decentralized and remotely conducted health studies have reported LTFU rates ranging from 18-30% [21–24]. To address this concern, detailed analysis was performed to identify the greatest sources of LTFU, which included evaluating the operational workflow and targeting specific sites with higher rates. All sites were also surveyed to determine where further support could be provided to improve participant retention. In response to our findings, the following strategies were implemented: 1) additional participant-facing materials were developed to serve as reminders and documentation of next steps in the study process; 2) site-specific support and reminders were provided to sites experiencing higher LTFU rates; 3) participant-specific flags were issued to the sites to prompt reminders and follow up calls to those participants noted to be LTFU; and 4) higher level of touch points were added for participants during at-home sample collection. As a result of these strategies, PD GENEration has achieved a LTFU rate of ∼6% for the current phase of the study.

Additionally, the <u>Parkinson’s Foundation Helpline</u> is a toll-free number for people with Parkinson’s, their families, friends and healthcare professionals to connect with a PD information specialist [25]. This resource is available in English and Spanish and allows the Foundation to be an information hub for the PD community. The resource has been particularly important for the functioning of the PD GENEration study. Staff members of the Helpline are trained on the study workflow and resources. As a result, they are equipped to answer questions related to genetic testing and counseling, clinical research, enrollment and blood draw logistics, and overall study processes. Since the beginning of the study, the Helpline has received 3,546 calls regarding PD GENEration, with 1,038 calls in 2025 alone.

## Data Acquisition and Management

As described above, all PD GENEration clinical data are captured through an EDC system by clinical research coordinators and site clinicians, or by participants themselves during virtual enrollment. API integrations maintained by the CRO and the Indiana University School of Medicine allow for the continuous integration and harmonization of data across the multiple platforms that support PD GENEration’s hybrid workflow. Processing of sample collection kits is managed by daily notifications regarding when a specific kit has been used to collect a sample, genetic reports are pulled directly from the genetic testing laboratory to share with clinicians, and patient information is accessible to central CGCs who provide remote results disclosures. Learnings from each phase of the study have informed improvements in data capture, resulting in the current system that focuses on collection of critical data, minimizes free text, and ensures appropriate checks regarding participant eligibility and study completion. The data science and management personnel of the PD GENEration team work closely with the data management team at the CRO to perform data cleaning and harmonization, ongoing data monitoring, and participant tracking to ensure quality of study data. Duplicate participants are also identified and consolidated using a matching algorithm based on combinations of participant birth date plus one or more of participant name, email address, and phone number. Internal workflows are in place to extract genetic testing results for internal analyses of variant positivity and frequency. For participants enrolling outside the US, data storage in a US-based EDC is addressed directly with the regulatory boards governing the individual participants’ consent during a country or site’s specific onboarding process, and consent language specific to the regulations around each site are added to ensure participants understand the data storage requirements.

## Data Sharing

In addition to returning genetic results to PwP, a central pillar of the study design has been the open sharing of data to advance discovery in PD risk and therapeutic development. At study onset, PD GENEration standardized the return of the raw genomic data from the clinical genetic testing laboratory, including variant calling format (VCF) and binary alignment map (BAM) files, to the Parkinson’s Foundation. These data are stored in a secure cloud-based system. Sites can request genomic data for their own participant cohort or can request study-wide de-identified data when necessary for site-specific research projects. All other data sharing is facilitated through the strategic partnership with ASAP and GP2. An established workflow is applied to de-identify and share all raw genomic data and demographic information captured by PD GENEration through GP2’s comprehensive data resource. As of December 2025, PD GENEration has released all collected clinical exome datasets and the first set of WGS datasets through this mechanism, with 14,686 and 2,008 datasets meeting quality standards for release, respectively. To access PD GENEration fully de-identified and uniquely coded genomic and demographic data released and governed by GP2, researchers must apply for <u>Tier 2 access</u> through Accelerating Medicines Partnership Parkinson’s Disease and Related Disorders (AMP-PD) and complete the GP2 <u>General Data Protection Regulation (GDPR) policy governed</u> <u>sample request form</u>.

The established partnership with GP2 also includes the sharing of residual DNA for long-term biobanking and future genomics analyses as approved by the PD GENEration Steering Committee. At this time, additional genomic analyses are only inclusive of a Neurobooster array to add to the compendium of data produced by GP2 largely for the purposes of large-scale genome-wide association studies (GWAS) [26].

## Budgetary Considerations

Budget and cost considerations have been an integral part of the study’s operational design. To ensure accessibility, the PD GENEration team determined at study onset that enrollment would be at no financial cost to participants. This meant that all financial costs directly related to study operations would be incurred by the Parkinson’s Foundation and its donors, which became an important factor in selecting vendors and partners to ensure best value without compromising expertise.

The primary factors driving per participant costs for PD GENEration were from the clinical genetic testing, which include CAP/CLIA certified genetic sequencing, clinical-grade variant curation and reporting, and data sharing and storage at the lab (57% of per participant cost), genetic counseling (8%), operational and data management through the CRO (16%), site and IRB costs (11%), and study logistics (8%). Study logistics include staffing costs, data storage, study material translation, patient recruitment, and engagement event fees. Including PD GENEration team staffing considerations, total per participant cost was approximately $1500 during early phases of the study using a clinical exome and rose to approximately $2700 following the transition to WGS and expanded site models. This increase reflects a more comprehensive sequencing backbone as well as the curation of the primary plus two additional gene panels.

## Results

As of March 31, 2026, PD GENEration has enrolled 32,577 participants from 10 countries, with 29,589 having completed genetic testing and counseling. Through our concentrated efforts to expand the study, we have doubled the enrollment achieved within the first 5 years of the study within only the last 20 months (**Figure 2**), with enrollment in the Registry: WGS Phase of the study (n = 16,574) now outpacing enrollment in the Pilot, Clinical, and Registry: Clinical Exome Phases combined (n = 16,003).

**Figure 2.**
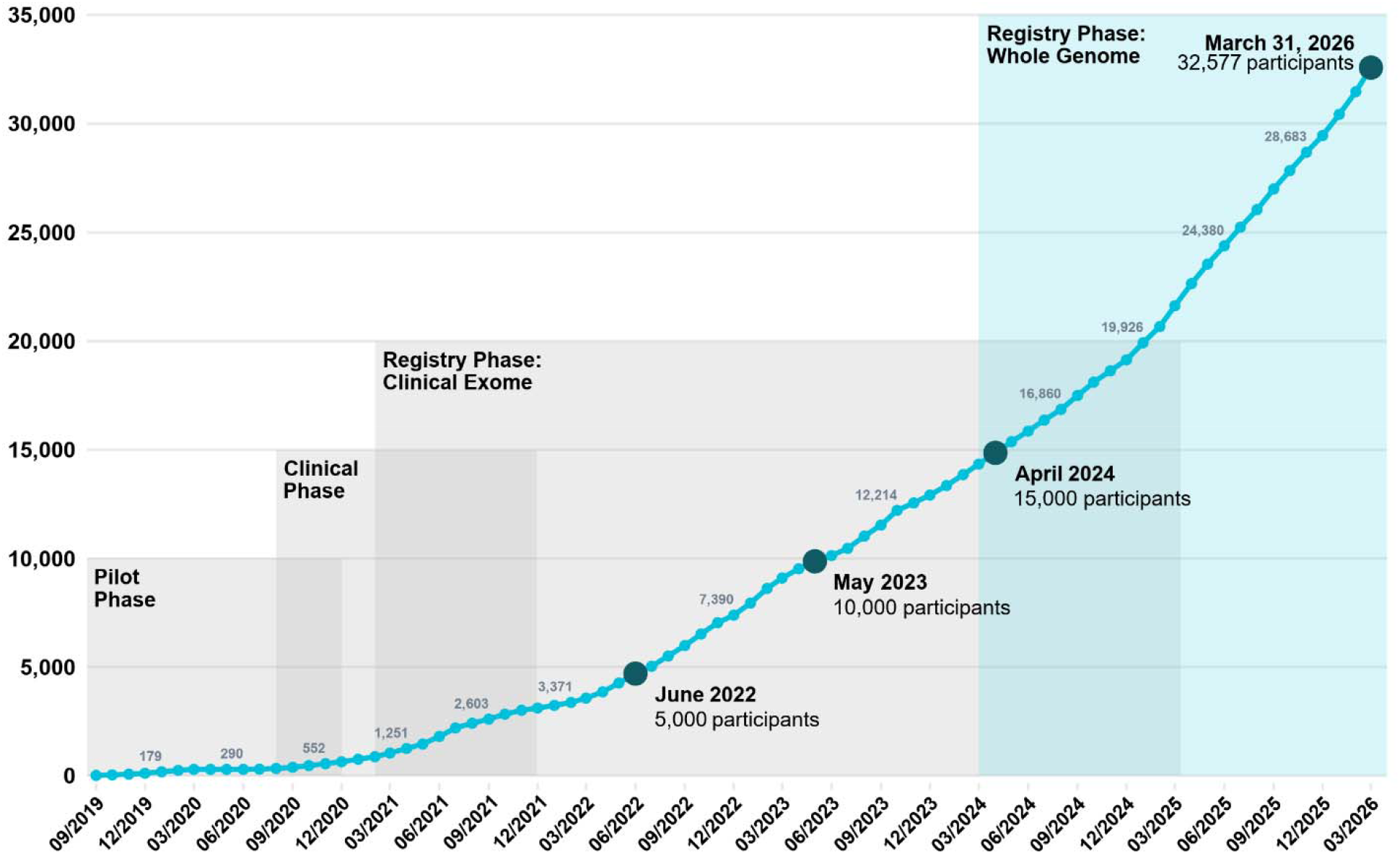
Cumulative enrollment of the PD GENEration study from September 2019 to March 2026. PD GENEration has enrolled over 32,000 participants clinically diagnosed with Parkinson’s disease (PD) through four study phases. The Pilot Phase assessed feasibility genetic testing on a backbone of clinical exome sequencing and return of results for people with Parkinson’s disease (PwP). The Clinical Phase marked a transition from a randomized pilot design to a broader implementation model with comprehensive clinical data collection. The Registry Phase: Clinical Exome further expanded the study to include three recruitment models while adopting a simplified protocol with limited clinical data collection. Finally, the Registry Phase: Whole Genome transitioned to using a backbone of WGS, began offering additional PD and non-PD related gene panels to participants and increased accessibility by expanding enrollment to countries across North, Central, and South America, the Caribbean, and Israel. Data presented includes all participants enrolled as of March 31, 2026.

Although most participants have been enrolled in the US, 3,265 (10.0%) have been enrolled at international sites throughout LATAM, Canada, and Israel (**Figure 3**). Of the 29,312 participants enrolled in the US, at least one participant has been enrolled in each of the 50 states (**Figure 4**). The Tasso+ kit has proven critical in expanding reach, with 52.9% of US-based participants using the kit (n = 4,614) to complete sample collection in 2025. Of these PwP, 875 (19.0%) lived in a state that does not currently have a PD GENEration local site and 602 (13.3% of those with residence data available) lived in a ZIP code classified as a “rural health area” by the Federal Office of Rural Health Policy [27]. Furthermore, 21.3% of participants enrolled in PD GENEration self-identified as belonging to racial and/or ethnic groups historically underrepresented in research (n = 6,941; **Figure 5**), and 74.5% reported having never previously participated in a PD-related research study (n = 22,351).

**Figure 3.**
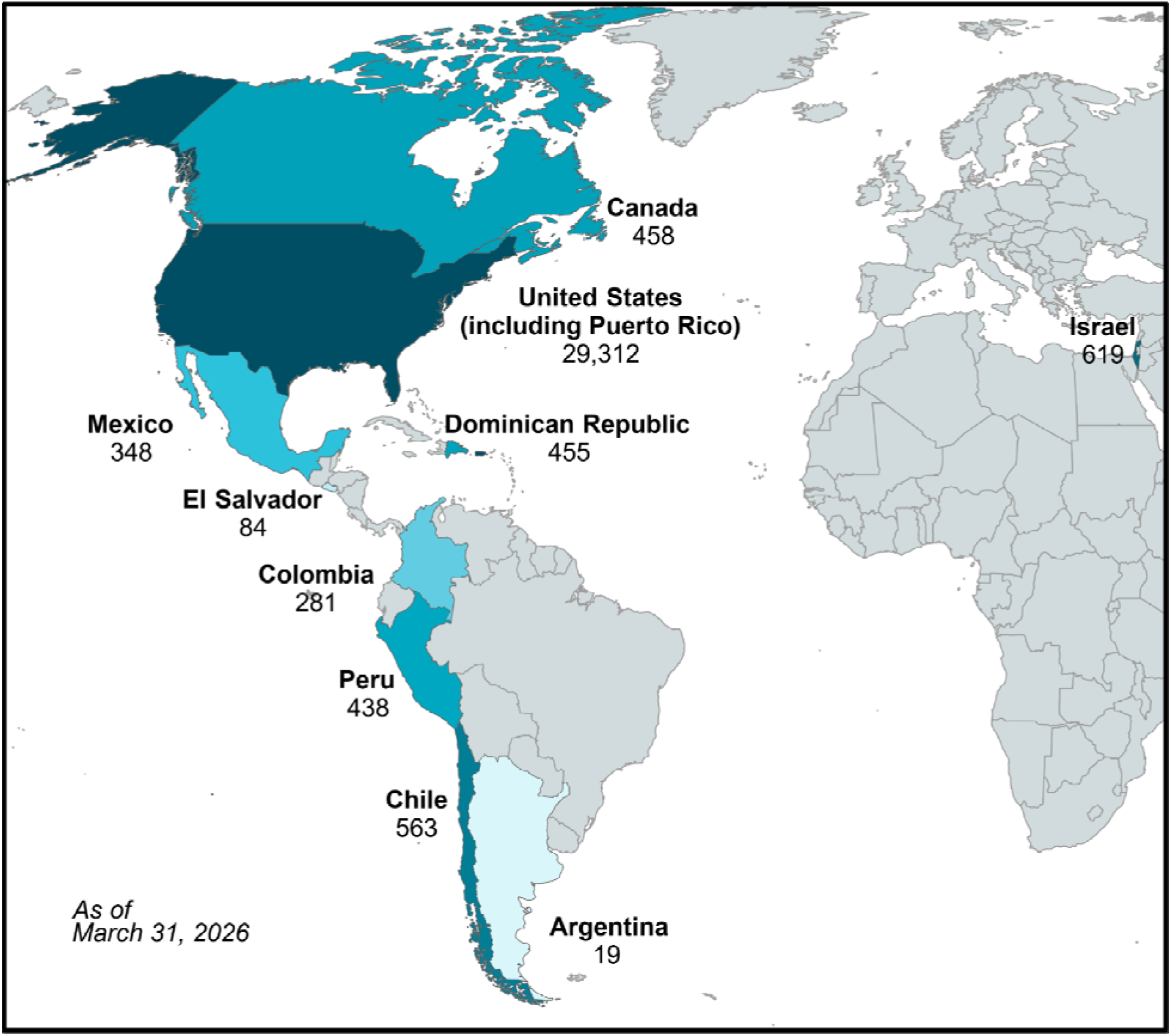
PD GENEration participant enrollment by country. Participant counts are based on the country in which the enrollment occurred. Data presented includes all participants enrolled as of March 31, 2026.

**Figure 4.**
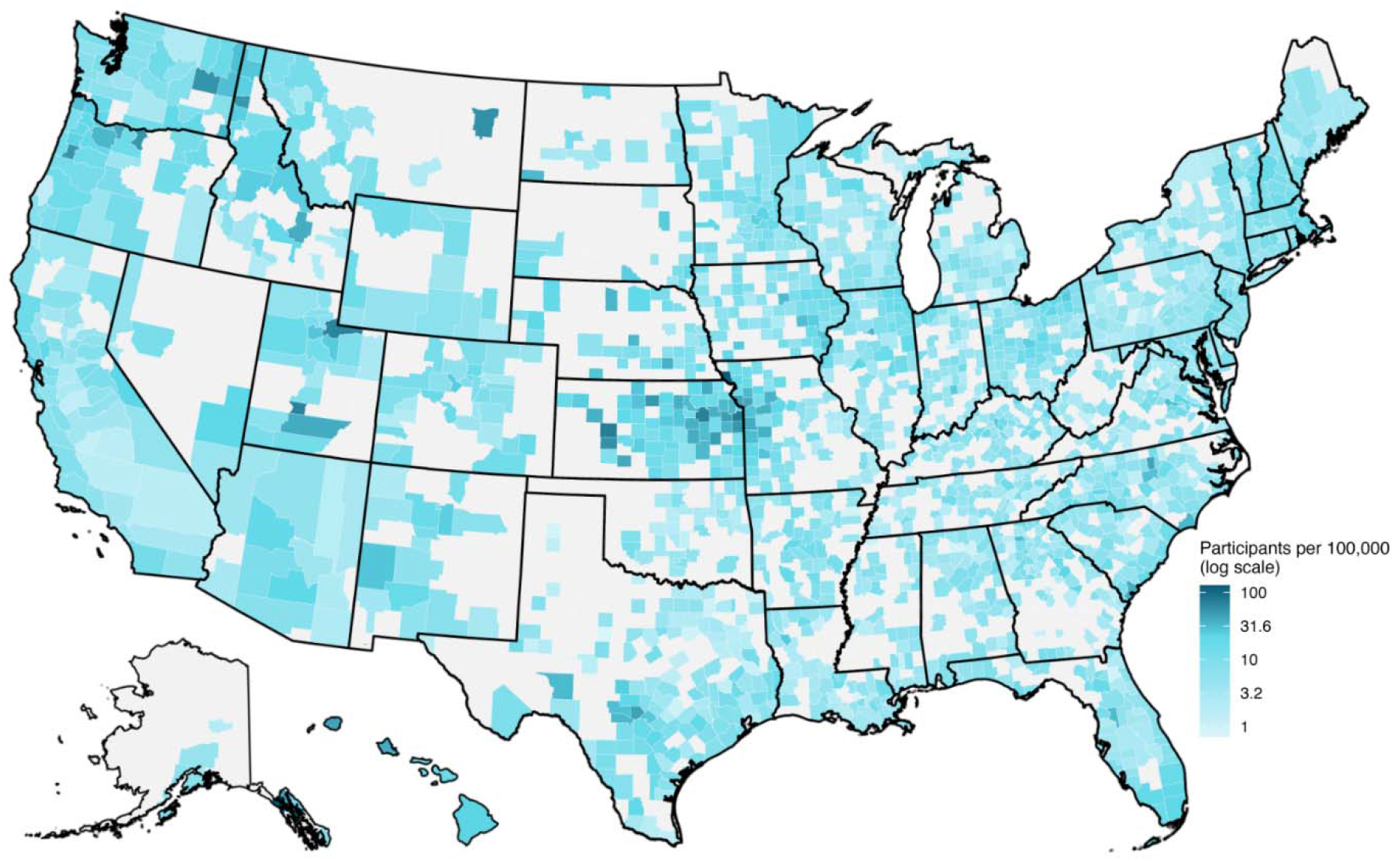
PD GENEration participant enrollment per capita by county in the United States. At least one person with Parkinson’s disease residing in each of the 50 states has enrolled in the study. Participant’s counties were defined using residence ZIP codes. Data presented includes all participants for which ZIP code data were available (n = 27,756) as of March 31, 2026.

**Figure 5.**
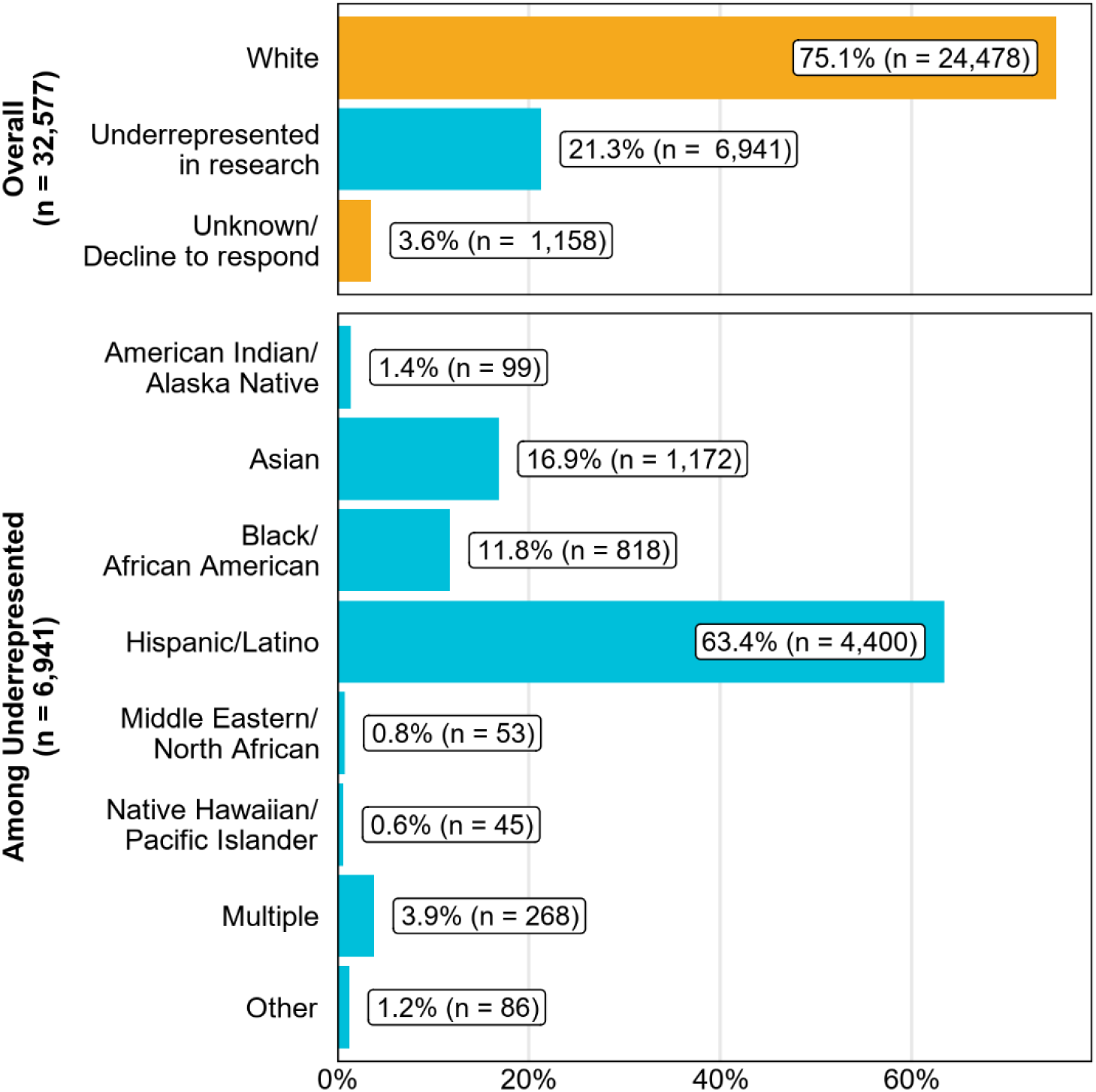
Self-reported race and ethnicity of people with Parkinson’s disease enrolled in PD GENEration. Of the 32,577 participants enrolled as of March 31, 2026, 21.3% self-identified as belonging to racial and/or ethnic groups historically underrepresented in research, including those that self-reported their race and/or ethnicity as American Indian/Alaska Native, Asian, Black/African American, Hispanic/Latino, Middle Eastern/North African, Native Hawaiian/Pacific Islander, Multiple, or Other.

Across all participants enrolled in PD GENEration, 59.1% reported their biological sex as male and 40.9% as female. The mean age of enrollment was 67.3 ± 10.6 years, mean age of PD onset was 60.2 ± 11.9 years, and mean age of PD diagnosis was 62.5 ± 11.2 years. Participants also provided general history of their PD, based on which 35.7% reported experiencing dyskinesia (10,691/29,949) and 7.9% reported having undergone deep brain stimulation surgery (2,550/32,165). Additionally, 20.1% reported an immediate family history of PD (6,317/31,432), defined as a first-degree relative having a confirmed diagnosis of PD. Complete participant demographics are outlined in (**Table 3**).

**Table 3.**
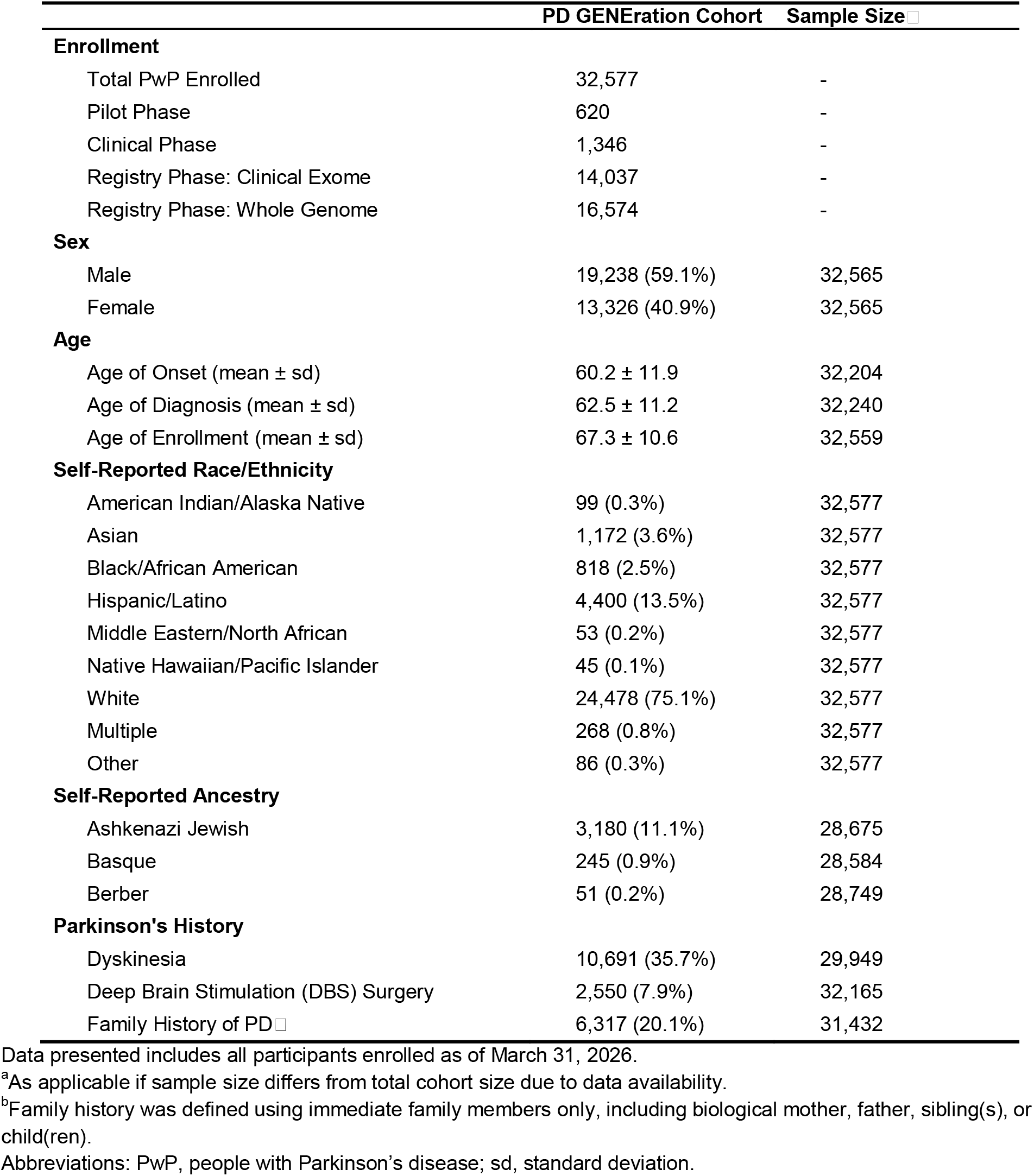
Demographics and Parkinson’s disease (PD)-related history of the participants enrolled in the PD GENEration study.

|  | PD GENERation Cohort | Sample Size<sup>□</sup> |
| --- | --- | --- |
| **Enrollment** |  |  |
| Total PwP Enrolled | 32,577 | - |
| Pilot Phase | 620 | - |
| Clinical Phase | 1,346 | - |
| Registry Phase: Clinical Exome | 14,037 | - |
| Registry Phase: Whole Genome | 16,574 | - |
| **Sex** |  |  |
| Male | 19,238 (59.1%) | 32,565 |
| Female | 13,326 (40.9%) | 32,565 |
| **Age** |  |  |
| Age of Onset (mean ± sd) | 60.2 ± 11.9 | 32,204 |
| Age of Diagnosis (mean ± sd) | 62.5 ± 11.2 | 32,240 |
| Age of Enrollment (mean ± sd) | 67.3 ± 10.6 | 32,559 |
| **Self-Reported Race/Ethnicity** |  |  |
| American Indian/Alaska Native | 99 (0.3%) | 32,577 |
| Asian | 1,172 (3.6%) | 32,577 |
| Black/African American | 818 (2.5%) | 32,577 |
| Hispanic/Latino | 4,400 (13.5%) | 32,577 |
| Middle Eastern/North African | 53 (0.2%) | 32,577 |
| Native Hawaiian/Pacific Islander | 45 (0.1%) | 32,577 |
| White | 24,478 (75.1%) | 32,577 |
| Multiple | 268 (0.8%) | 32,577 |
| Other | 86 (0.3%) | 32,577 |
| **Self-Reported Ancestry** |  |  |
| Ashkenazi Jewish | 3,180 (11.1%) | 28,675 |
| Basque | 245 (0.9%) | 28,584 |
| Berber | 51 (0.2%) | 28,749 |
| **Parkinson's History** |  |  |
| Dyskinesia | 10,691 (35.7%) | 29,949 |
| Deep Brain Stimulation (DBS) Surgery | 2,550 (7.9%) | 32,165 |
| Family History of PD<sup>□</sup> | 6,317 (20.1%) | 31,432 |

Data presented includes all participants enrolled as of March 31, 2026.
<sup>a</sup>As applicable if sample size differs from total cohort size due to data availability.
<sup>b</sup>Family history was defined using immediate family members only, including biological mother, father, sibling(s), or child(ren).
Abbreviations: PwP, people with Parkinson's disease; sd, standard deviation.

As of March 31, 2026, primary PD panel genetic testing was completed for 15,169 participants enrolled in the Pilot, Clinical, and Registry: Clinical Exome Phases of the study and 14,420 participants enrolled in the Registry: WGS Phase of the study. We observed a cumulative genetic positivity rate of 12.1% amongst these PwP, representing participants that carried at least one likely pathogenic or pathogenic variant in the primary panel genes. Further 9,831 PD GENEration participants completed secondary PD panel genetic testing and 10,120 completed CDC 10 Tier 1 panel genetic testing, with genetic positivity rates of 4.4% and 1.5%, respectively. Comprehensive analyses of the genetic landscape identified in the cohort, including variant-level analyses and genotype-phenotype correlations are ongoing and will be presented in future dedicated genetics reports.

## Discussion

PD GENEration represents a large-scale, international effort to expand access to CAP/CLIA certified genetic testing and counseling for PwP while also supporting research into the genetic architecture of PD. The study demonstrates the feasibility of integrating genetic testing into clinical and research settings and that the implementation of multiple recruitment models enables broader participation across diverse populations and geographic regions. These approaches highlight the potential for scalable and accessible genetic testing programs in PD research.

## Accessibility Driving Operational Design

The operational and strategic design of PD GENEration has pivoted several times over the past six years to meet one of the main priorities of the study—to make genetic testing and counseling more accessible to PwP. Accessibility efforts have focused on reducing barriers to participation, expanding opportunities for enrollment, and ensuring the study is inclusive and accessible to a broad and diverse participant population. Several different considerations and strategies were implemented to uphold these standards.

### Reducing participant and site burden

Although the Pilot and Clinical Phases of PD GENEration showed the acceptability of the study amongst participants and the research community was reported with high satisfaction [11], anecdotal feedback from the enrolling clinicians and coordinators demonstrated that the extensive clinical data collection was burdensome, limiting the number of enrolled participants. There was also concern regarding participant strain, especially for those who were new to clinical research or were historically marginalized or mistreated in research settings. Although extended surveys and scales were useful for knowledge advancement, the study pivoted to prioritize a more balanced approach between effective genetic testing and the needs of the community. With the goal of enrolling 15,000 participants, the streamlined Registry Phase was launched, focusing on expanding outreach to PwP and supporting more sites was deemed more economically and structurally beneficial than maintaining the extended clinical data collection. To accomplish the high enrollment goal, the study expanded access through the use of virtual enrollment.

### Virtual enrollment

The nationally enrolling supersite model improves study access by enabling virtual enrollment for participants across the US, regardless of geographic location, which is further aided by the study’s partnership with Tasso+ that allows for at-home blood draws, as described in the Study Workflow. The success of this model was demonstrated by the increase of widespread enrollment observed during the Clinical Exome Registry and WGS Registry Phases, as well as the wide-spread use of the Tasso+ device for blood collection in 2025. The model has resulted in improved outreach to new participants of diverse backgrounds and has allowed PwP from rural health areas that may not have access to traditional clinical trial centers to participate in research.

Although the virtual model has shown success in expanding enrollment, it did not come without barriers and challenges. The identification of supersites that could handle increased capability to enroll participants from any state required extensive IRB, staffing, time zone, and cost considerations. We also had to identify an appropriate solution to allow for at-home blood draws, which led to the partnership with Tasso+. Finally, we needed to determine how to engage these participants. Leveraging the established work of the Parkinson’s Foundation in communities throughout the US and through establishing the Referral site model, we were able to see the impact of the combination of extensive outreach and unique operations coming together. Overcoming these barriers has enabled broader access to genetic testing and counseling, thereby extending the impact of research across the PD community.

### Language expansion

Based on recent Census data, 20.0% of people in the US identify as Hispanic/Latino [28], and 13.3% of people in the US speak Spanish at home [29]. The high prevalence of PD throughout the US highlights the importance of making the study accessible to as many potential participants as possible, regardless of language. Therefore, all participant-facing documents have been made available in both English and Spanish, including recruitment materials, informed consent forms, Tasso+ instructional videos, genetic counseling fact sheets, and community-based presentations.

The study has also engaged referral sites and local sites that predominantly provide care to Spanish-speaking participants and has established supersites with Spanish-speaking coordinators, clinicians, and CGCs to provide better accessibility to the study in the US. We aim to continue onboarding additional international sites and expand language capabilities to accommodate more participant enrollment from non-English or Spanish speakers. For example, with current plans for expansion into Brazil and Québec, Canada, Brazilian Portuguese and French-Canadian materials are being developed to accommodate the local needs of the sites and the patient community.

### Underrepresented populations

It is well-established that a lack of ancestral diversity persists in the landscape of PD genetic research [30]. Yet, more comprehensive analyses of diverse populations remain critical as it is unclear whether our current understanding of genetic contributors to PD are relevant across ancestral populations. We also must consider whether population-specific variation may exist such as the Colombian *GBA1* variant (p.K198E) or the recently described *GBA1* intronic risk factor variant (rs3115534-G) identified in ∼50% of PwP from West Africa [31, 32]. Therefore, one of the central goals of PD GENEration is to ensure the study is accessible to all PwP, including those historically underrepresented in clinical trials and PD genetics research. This objective is further supported by the study’s partnership with ASAP and GP2, ensuring that the PD GENEration cohort contributing to GP2’s global genetics effort reflects the real-world population of individuals living with PD.

Toward this goal, PD GENEration implemented several strategies focused on building trust and engagement with underrepresented populations, such as partnering with new local and referral sites that serve these communities. The study also prioritized diverse enrollment across all sites and provided resources to support educational and enrollment opportunities within these populations. For example, in partnership with GP2, PD GENEration granted support to 20 local and referral sites to specifically aid in their engagement of underrepresented populations. In turn, site staff were able to attend more support groups, discuss research and genetics through participant symposiums, and increase the accessibility of PD GENEration to more diverse populations by introducing testing to new communities. Additionally, the PD GENEration team planned and executed study-focused recruitment events at existing support groups, community events, and educational events in the US and internationally, primarily focusing on underrepresented populations in research. Partnerships with other groups supporting underrepresented populations in PD research, such as LARGE-PD, HAWAII-PD, and Black and African American Connections to PD (BLAAC-PD), have also been established.

By implementing these strategies over the last 2 years, the study has seen an increase in participation of PwP who identify as belonging to an underrepresented population based on race and ethnicity, rising from 15.7% at the beginning of 2024 to 21.3% at present. PD GENEration continues its commitment to ensuring the accessibility of the study and prioritizing the needs of underrepresented communities as it continues to grow.

## Feasibility and Sustainability

Throughout its iterations, PD GENEration has pivoted to consider the needs of the PD and research communities. With new phases came new sequencing and operational considerations that altered the cost per participant to meet the needs of the changing ecosystem of PD genetics. The PD GENEration team continuously evaluates the cost per participant ensuring that with the budget allocated, the study can enroll as many PwP as possible. Cost-lowering initiatives have included re-negotiating laboratory costs based on volume, creating standard operating procedures with the CRO for site metrics and reporting to limit excessive operations costs, and lowering LTFU rates to ensure study completion. However, the cost of providing CAP/CLIA-accredited genetic testing, genetic counseling, and operations of a study at this scale remains prohibitive and requires a significant budget. Nevertheless, based upon the steady decline in sequencing costs [33], we anticipate a reduction in the per participant costs in the years to come.

Importantly, the study costs support a participant-centric model, as outlined here, that ensures accessibility, clinical relevance and research value, appropriate disclosure of results, and sustained engagement, all while maintaining fiscal responsibility. This model was supportable for a study of this scale for the PD community; however, there are several areas within the PD GENEration infrastructure that can be remodeled to fit different budgets and needs of different disease indications, such as the type of sequencing coverage, the number of different genetic testing panels returned, the services contracted for with the CRO and genetic counseling cores, and/or the need for translation of participant-facing documents. Yet, until genetic testing in PD becomes the standard of care with consistent reimbursement through insurance providers, a study of this nature requires substantial resources.

## Contributions to Research

Beyond the direct benefit to PwP and to their clinical care that is provided through genetic testing, PD GENEration actively contributes to PD research via the distribution of de-identified raw genomic sequencing data through partnership with GP2. These data provide opportunities for researchers globally to improve the understanding of PD genetic architecture and molecular mechanisms, identify novel genetic factors that may inform therapeutic development, and expand knowledge of PD genetic risk in populations historically underrepresented in clinical research.

Importantly, PD GENEration was designed to integrate these research objectives with a clinical genetic testing infrastructure. Testing through a CAP/CLIA-accredited laboratory provides a framework for clinical interpretation and variant reporting that meets regulatory and quality requirements for clinical laboratory testing and clinical reporting to participants. This is relevant in the context of genetic research, where typically research-grade findings do not have sufficient evidence to establish the clinical validity necessary for return of results [34]. Maintaining the clinical testing infrastructure alongside the study’s research objectives and partnership with GP2 is aimed at fostering new discoveries that can further the interpretation of genetic findings identified in PD GENEration participants. For example, as GP2 generates additional evidence regarding the contributions of specific genes and variants to PD, this may support the re-evaluation of previously identified results and, when appropriate, the return of those results to participants. Therefore, the clinical and research components of PD GENEration are complementary, as advances in PD genetics are aimed to benefit the individuals who are contributing to the study. The synergy provides a pathway for translating genetic research into clinical knowledge, and ultimately, advancing precision medicine efforts in PD.

## Future Directions

### PD GENEration Sub-Studies

PD GENEration has continuously evolved based on the needs of the PD and research community. Consistently, participants are looking for additional opportunities to contribute to research [11], and researchers are requesting information to supplement the genetic data being collected. In response, PD GENEration created three new pilot sub-studies in 2025 (**Supplemental Figure 1**). First, PD GENEration Surveys periodically deploys retrospective and cross-sectional surveys to the entire cohort on topics of interest from the research community. The first survey was completed in September 2025. Next, PD GENEration Family offers genetic testing and counseling at no-cost to first-degree family members of select PD GENEration participants, including parents, siblings, and adult children. Launched in October 2025, the sub-study supports the identification of at-risk individuals and helps link family member data together to better understand genetic inheritance and penetrance of PD within families. Finally, PD GENEration Insights is a longitudinal research study set to launch in May 2026 and collect extensive phenotypic data from select PD GENEration participants to learn more about the clinical presentation and progression of genetic forms of PD.

### Participant Engagement

PD GENEration is developing a participant-facing component to the study’s EDC system, essentially acting as a participant portal to further promote participant retention and provide the opportunity for participants to engage with additional PD GENEration research studies, as discussed above. The participant “portal” will also allow the PD GENEration team to actively keep the study population engaged, providing participants with real-time updates on their status in the study, sharing study-wide results, informing participants of additional research opportunities, and providing educational materials about PD and clinical research. Based on current projects, we anticipate the portal launch to occur by Autumn 2026.

## Conclusion

The Parkinson’s Foundation through PD GENEration has established a large-scale infrastructure that has enabled genetic testing of more than 32,000 PwP across ten countries. The study protocol and methods have evolved to reflect evolution of science and to be responsive to the patient and research community’s needs. Together, these insights and our framework provide a model for scaling clinical genetic testing initiatives across diverse research and clinical populations.

## Supporting information

The Parkinson’s Foundation PD GENEration Study

## Data Availability

Access to PD GENEration genomic data released through the Global Parkinson’s Genetics Program (GP2) requires researchers to obtain Tier 2 access through the Accelerating Medicines Partnership Parkinson’s Disease and Related Disorders (AMP-PD) platform and to complete the GP2 General Data Protection Regulation (GDPR) policy governed sample request process.

https://amp-pdrd.org/register-for-amp-pd

## Declarations

### List of Abbreviations

ACMG: American College of Medical Genetics and Genomics
AMP: Association for Molecular Pathology
AMP-PD: Accelerating Medicines Partnership Parkinson’s Disease and Related Disorders
API: Application programming interface
ASAP: Aligning Science Across Parkinson’s
BAM: Binary alignment map
BLAAC-PD: Black and African American Connections to Parkinson’s Disease
CAP: College of American Pathologists
CLIA: Clinical Laboratory Improvement Amendments
CRO: Contract Research Organization
GDPR: General Data Protection Regulation
FDA: Food and Drug Administration
GP2: Global Parkinson’s Genetics Program
GWAS: Genome-wide association studies
IRB: Institutional Review Board
LAR: Legally authorized representative
LARGE-PD: Latin American Research Consortium on the Genetics of Parkinson’s Disease
LATAM: Latin America (Argentina, Chile, Colombia, El Salvador, Dominican Republic, Mexico, and Peru)
LTFU: Lost to follow-up
MDS: Movement Disorder Society
MDS-UPDRS: Movement Disorder Society–Unified Parkinson’s Disease Rating Scale
MoCA: Montreal Cognitive Assessment
PD: Parkinson’s disease
POA: Power of attorney
PPAC: People with Parkinson’s Advisory Council PSG Parkinson Study Group
PwP: People with Parkinson’s
US: United States
VCF: Variant call format
WGS: Whole-genome sequencing

## Ethics Approval and Consent

This study was conducted in accordance with applicable ethical and regulatory requirements and received approval from a central institutional review board, Advarra, Inc., as well as local site-based institutional review boards/ethics committees, as applicable. Written informed consent was obtained from all participants prior to participation in the study.

## Consent for Publication

Not applicable.

## Competing Interests

K.G.G. is an employee of the Parkinson’s Foundation.

A.A.D. is a contractor for the Parkinson’s Foundation.

M.D. is an employee of the Parkinson’s Foundation.

R.D.L. is an employee of the Parkinson’s Foundation.

M.T. is an employee of the Parkinson’s Foundation.

I.A. is an employee of the Parkinson’s Foundation.

N.B. is an employee of the Parkinson’s Foundation.

L.C. is an employee of the Parkinson’s Foundation.

A.C.Z. is an employee of the Parkinson’s Foundation.

K.D. is an employee of the Parkinson’s Foundation.

M.F. is an employee of the Parkinson’s Foundation.

M.Nicewaner is an employee of the Parkinson’s Foundation.

S.O. is an employee of the Parkinson’s Foundation.

J.R. is an employee of the Parkinson’s Foundation.

A.Y. is an employee of the Parkinson’s Foundation.

A.D. is an employee of the Parkinson’s Foundation.

T.F. has received research funding from the Parkinson’s Foundation, the Michael J. Fox Foundation for Parkinson’s Research National Institutes of Health and serves on the external advisory board on several Alzheimer’s Disease Research Centers.

A.H. has received honoraria for her patient research advocacy from the Parkinson’s Foundation (member of the PD GENEration Steering Committee), Duke University (member of the joint U.S. Federal Drug Administration and Clinical Trials Transformation Initiative Patient Engagement Collaborative) and Novartis (member of Novartis’s PEATT global patient engagement initiative).

L.H. is supported by research funding from the Parkinson’s Foundation and Michael J Fox Foundation for Parkinson’s Research.

S.W.L. is employed by Navitas Clinical Research, the CRO that is supporting PD GENEration.

K.M. has no conflict of interest to disclose.

I.M. has received research funding from the Parkinson’s Foundation, Michael J. Fox Foundation for Parkinson’s Research, Aligning Science Across Parkinson’s Global Parkinson’s Genetic Program (ASAP-GP2), National Institutes of Health, Veterans Affairs Health Care System and American Parkinson’s Disease Association. He is also a member of PD GENEration’s Steering Committee and GP2 Operations Committee.

N.E.M. is supported by the National Institute of Health (grant 1K08NS131581) and the Align Science Across Parkinson’s (ASAP) Global Parkinson’s Genetics Program (GP2). He is a member of the steering committee of the PD GENEration study for which he receives an honorarium from the Parkinson’s Foundation.

A.N. has no conflict of interest to disclose.

M.Nance has received research funding from the Parkinson’s Foundation, Bial, Novartis, and CHDI Foundation; Center of Excellence funding from the Parkinson Foundation and the Huntington Disease Society of America; compensation for activities on behalf of the Huntington Study Group and the Parkinson Study Group, personal compensation for consulting activities on behalf of Arrowhead Pharmaceuticals, Roche, and Uniqure. She serves on the PD GENEration Steering Committee. She serves on the Clinical Care Committee of the Huntington Study Group.

J.P. is a member of the Parkinson’s Foundation People with Parkinson’s Foundation Advisory Council, President of the Mid-Atlantic Chapter of the Parkinson’s Foundation, a member of the ABIM Sleep Advisory Governing Committee, and a member of the NAPS Community Engagement Board.

R.B.S. has received research funding from the Parkinson’s Foundation, Michael J. Fox Foundation for Parkinson’s Research, National Institutes of Health, Bial, Acadia

Pharmaceuticals, and CHDI Foundation, and personal compensation for participation on Data Safety and Monitoring Boards for HEALEY ALS, AskBio, and Appello.

M.A.S. has received research funding from the Parkinson’s Foundation, Michael J. Fox Foundation for Parkinson’s Research, National Institutes of Health, Cure Parkinson’s, Farmer Family Foundation, US FDA, and personal compensation for advisory or steering committee services from Northwestern Univ., Sutter Health, SPARK NS, and the Parkinson Study Group (for its services to Biogen and UCB Pharma/Novartis).

T.S. has served as a consultant for Blue Rock Therapeutics, Centessa, Critical Path for Parkinson’s Consortium (CPP), MJFF, Prevail/Lilly, Roche/Genentech, Ventus, Sinopia, Takeda, Vanqua Bio, Ventus, Ventyx, and VIMA Tx. T.S. has equity in Sinopia and has served on the ad board for AskBio, Biohaven, Booster, GAIN, Janssen, Neuron23, Novartis, Parkinson Study Group, Prevail/Lilly, and Roche/Genentech. T.S. has served as a member of the scientific advisory board of Koneksa and UCB. T.S. has received research funding from Neuroderm, Prevail, Roche, NINDS, MJFF, and Parkinson’s Foundation.

J.V. is supported by research funding from the Parkinson’s Foundation and Michael J Fox Foundation for Parkinson’s Research.

A.M.W. has research funding from NIH/NIH Award R44AG080861 and NIA/NIH Award R01AG085029, from the Parkinson’s Foundation, and has participated in clinical trials funded by Roche/Genentech, Biogen/Denali, Bial, Amylyx, Ferrer, Ono, and Biohaven. She has received consultant payments from Accordant, CVS/Caremark, Genentech, Amylyx and Ono Pharmaceuticals.

Y.L. is employed by Navitas Clinical Research, the CRO that is supporting PD GENEration

H.G. is an employee and shareholder of Fulgent Genetics.

B.C. is an employee of Tasso Inc.

C.B. has no conflict of interest to disclose.

A.B.S. works as a contractor for the Global Parkinson’s Genetics Program and receives grant support from the Michael J Fox Foundation for Parkinson’s Research.

J.C.B. is an employee of the Parkinson’s Foundation.

R.N.A. has research that is funded by the Michael J. Fox Foundation, the Silverstein Foundation, the Parkinson’s Foundation and the Aufzien Family Center for the Prevention and Treatment of Parkinson’s Disease. He received consultation fees from Bexxion, Biogen, Biohaven, Capsida, Gain Therapeutics, Genzyme/Sanofi, Janssen, SK Biopharmaceuticals, Takeda, Teva and Vanqua Bio.

## Funding

PD GENEration is sponsored by the Parkinson’s Foundation in partnership with Aligning Science Across Parkinson’s (ASAP) through the Global Parkinson’s Genetics Program (GP2).

## Author Contributions

Study leadership and supervision: K.G.G., J.C.B., R.N.A.

Past study leadership: A.N.

Study operations: K.G.G., A.A.D., M.D., R.D.L., M.T., I.A., N.A., L.C., A.C.Z., K.D., M.F.,

M.Nicewaner, S.O., J.R., A.Y., L.H., S.W.L., Y.L., J.C.B.

Study steering committee: T.F., K.G.G., A.H., L.H., K.M., I.M., N.E.M., A.N., M.Nance, J.P.,

R.B.S., M.A.S., T.S., J.V., A.M.W., J.C.B., R.N.A.

Genetic counseling: T.F., L.H., J.V.

Laboratory resources and sequencing support: H.G., B.C. Formal analysis: K.G.G., A.A.D., M.T., L.C.

Visualization: A.A.D., M.T., L.C.

Writing – original draft: K.G.G., A.A.D., M.D., R.D.L., M.T., I.A., N.B., L.C., A.C.Z., K.D., M.F., M.Nicewaner, S.O., J.R., A.Y., J.C.B.

Writing – review & editing: all authors.

All authors have reviewed and approved the final manuscript.

## Acknowledgements

The Parkinson’s Foundation PD GENEration study is a flagship initiative of the Parkinson’s Foundation in partnership with the Global Parkinson’s Genetics Program (GP2), a resource program of the Aligning Science Across Parkinson’s (ASAP) initiative. Additional financial and in-kind support comes from the Parkinson’s community – industry partners, nonprofit organizations, and individuals whose lives have been touched by Parkinson’s. For up-to-date information on the study, visit https://www.parkinson.org/pdgeneration. The Parkinson’s Foundation and the PD GENEration authors are indebted to the support, interest, and participation of those living with Parkinson’s disease without which this study would not be possible.

## Notes

### Summary of Updates

The revised manuscript incorporates revisions following peer review during submission to a journal. The manuscript has been substantially reorganized, with expanded detail on PD GENEration operations. New and expanded content addresses barriers and facilitators, sustainability, infrastructure requirements, and costs. The manuscript has also been clarified and strengthened throughout.

