## Supplemental for "PD GENEration: An International Parkinson’s Disease Genetic Research Study"

**Supplemental Table 1. PD GENEration Registry Phase: Whole Genome Sequencing study inclusion and exclusion criteria.**

| Study Criteria | Details |
|---|---|
| **Inclusion Criteria** | **Study Population**: People With Parkinson's disease (PWP)<br><br>1. Meet Movement Disorder Society (MDS) Clinical Diagnostic Criteria for Parkinson's disease: probable diagnosis based on Investigator discretion.<br>2. Willingness to undergo genetic testing and may choose to be informed of genetic test results for, at minimum, seven Parkinson's-related genes, including: *GBA1, LRRK2, SNCA, VPS35, PRKN, PINK1, PARK7*. Participants may choose to also receive additional findings which might be related to their PD diagnosis and/or health-related clinically actionable findings.<br>3. Based on site clinician's determination, have the capacity to give full informed consent in writing or electronically, or provide consent through a legally authorized representative (LAR)/ power of attorney (POA), and have read, understood and completed the informed consent form.<br>4. Can perform, or have a designee who can perform, study activities (including completion of either online, in-person, or paper surveys). |
| **Exclusion Criteria** | 1. Probable diagnosis at the time of consent of an atypical parkinsonian disorder (i.e., multiple system atrophy, progressive supranuclear palsy, dementia with Lewy bodies, corticobasal syndrome), including that due to medications, metabolic disorders, encephalitis, cerebrovascular disease, or normal pressure hydrocephalus<br>2. Individuals who have received a blood transfusion within the past 3 months.<br>3. Individuals who have active hematologic malignancies such as lymphoma or leukemia.<br>4. Individuals who have had a bone marrow transplant.<br>5. Individuals under the age of 18 or age of majority in applicable states at the time of consenting |

Study inclusion and exclusion criteria are accurate as of March 31, 2026.

**Supplemental Table 2. PD GENEration local sites and supersites per country actively enrolling participants as of March 31, 2026.**

| Country | Active Local Sites (n) | Active Supersites (n) |
|---|---|---|
| Argentina | 1 | - |
| Canada | 2 | - |
| Chile | 4 | - |
| Colombia | 1 | - |
| Dominican Republic | 1 | - |
| El Salvador | 1 | - |
| Israel | 3 | - |
| Mexico | 3 | - |
| Peru | 1 | - |
| United States | 62 | 6 |
| **Total** | **79** | **6** |

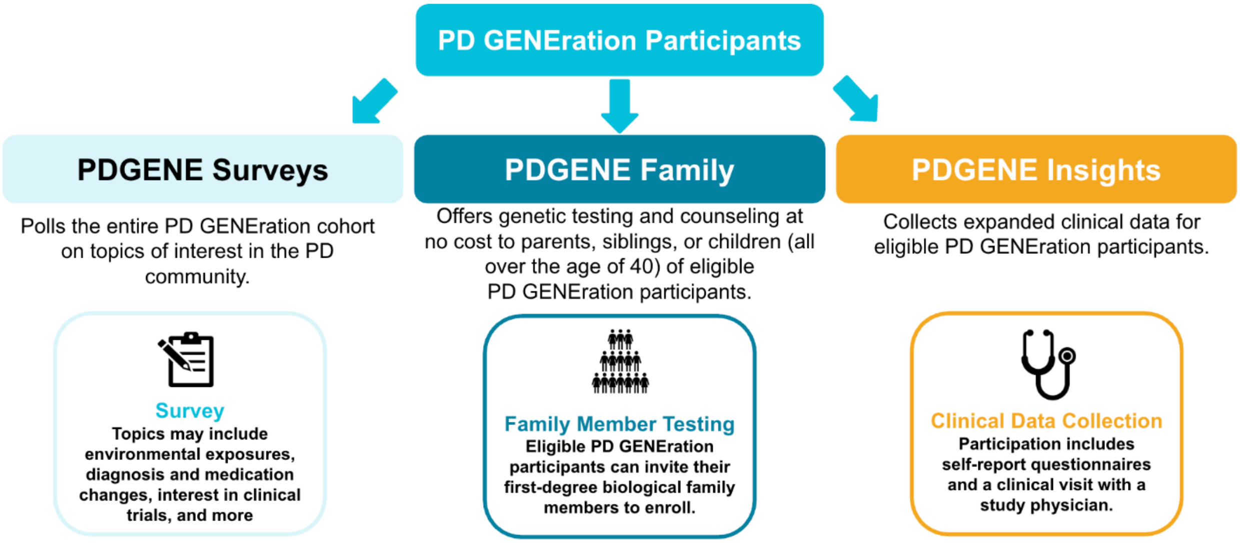


**Supplemental Figure 1. PD GENEration sub-studies available to select participants that have completed the core PD GENEration study workflow.**
